# Whole Genome Rare-Variant Association Study of HIV-1 Progression in a Southern African Population

**DOI:** 10.1101/2020.12.16.20248307

**Authors:** Prisca K. Thami, Wonderful Choga, Delesa D. Mulisa, Collet Dandara, Andrey K. Shevchenko, Melvin M. Leteane, Vlad Novitsky, Stephen J. O’Brien, Myron Essex, Simani Gaseitsiwe, Emile R. Chimusa

**Affiliations:** Division of Human Genetics, Department of Pathology, Institute of Infectious Disease and Molecular Medicine, University of Cape Town, Cape Town, 7925, South Africa; Botswana Harvard AIDS Institute Partnership, Gaborone, Botswana; Theodosius Dobzhansky Center for Genome Bioinformatics, St. Petersburg State University, St. Petersburg 199034, Russia; Department of Biological Sciences, University of Botswana, Gaborone, Botswana; Harvard T.H. Chan School of Public Health AIDS Initiative, Department of Immunology and Infectious Diseases, Harvard T.H. Chan School of Public Health, Boston, Massachusetts, 02115, USA; Laboratory of Genomics Diversity, Center for Computer Technologies, ITMO University, St. Petersburg, 197101, Russia; Guy Harvey Oceanographic Center Halmos College of Natural Sciences and Oceanography Nova Southeastern University, Ft Lauderdale, Florida, 33004, USA

**Author notes:** **Corresponding author:** Emile R. Chimusa, Division of Human Genetics, Department of Pathology, Institute of Infectious Disease and Molecular Medicine, University of Cape Town, Anzio Road, Observatory, 7925, Cape Town, South Africa.

**Keywords:** Rare-variant Association, Whole Genome Sequencing, host genetics, HIV-1, progression

## Abstract

Despite the high burden of HIV-1 in Botswana, the population of Botswana is significantly underrepresentation in host genetics studies of HIV-1. Furthermore, the bulk of previous genomics studies evaluated common human genetic variations, however, there is increasing evidence of the influence of rare variants in the outcome of diseases which may be uncovered by comprehensive complete and deep genome sequencing. This research aimed to evaluate the role of rare-variants in susceptibility to HIV-1 and progression through whole genome sequencing. Whole genome sequences (WGS) of 265 HIV-1 positive and 125 were HIV-1 negative unrelated individuals from Botswana were mapped to the human reference genome GRCh38. Population joint variant calling was performed using Genome Analysis Tool Kit (GATK) and BCFTools. Cumulative effects of rare variant sets on susceptibility to HIV-1 and progression (CD4+ T-cell decline) were determined with optimized Sequence Kernel Association Test (SKAT-O). *In silico* functional analysis of the prioritized variants was performed through gene-set enrichment using databases in GeneMANIA and Enrichr. Novel rare-variants within the *ANKRD39* (8.48 × 10^−8^), *LOC105378523* (7.45 × 10^−7^) and *GTF3C3* (1.36 × 10^−6^) genes were significantly associated with HIV-1 progression. Functional analysis revealed that these genes are involved in viral translation and transcription. These findings highlight the significance of whole genome sequencing in pinpointing rare-variants of clinical relevance. The research contributes towards a deeper understanding of the host genetics HIV-1 and offers promise of population specific interventions against HIV-1.

## INTRODUCTION

Human immunodeficiency virus (HIV) infection remains one of the world’s worst pandemics. Over 37 million people lived with HIV by the end of 2018 globally. About half of these live in Southern and eastern Africa. Botswana is the third most affected country in Southern Africa, after eSwatini and South Africa in first and second positions respectively (1). The country is affected predominantly by HIV-1C. The HIV epidemic became severe in Botswana by the late 1990s at a prevalence of 30-40% in pregnant women (2).

Botswana was the first country in Southern Africa to offer free antiretroviral therapy (ART) to people infected with HIV. Due to the rapid scaleup of anti-HIV drugs, there has been a sharp decline in HIV-related morbidity and mortality (3,4). HIV prevalence in Botswana has since lowered to 20.3% among adults (1). Nonetheless Botswana still remains one of the most affected countries globally due to the high baseline HIV prevalence and a successful national ART programme.

There is a remarkable interpersonal heterogeneity of HIV-1 phenotypes (acquisition, progression and drug metabolism). This heterogeneity in host phenotypes of HIV-1 infection has been attributed to several factors including host genetics (5–7). Treatment against HIV-1 does not offer cure and to date no effective vaccine has been found against HIV infection (8,9). Therefore, identifying population specific genetic factors can catapult the invention of effective strategies against HIV-1 in African populations.

Candidate disease gene methods revealed a number of genes associated with HIV-1 infection (10–12). A momentous achievement in the HIV-1 candidate gene research was the discovery of *CCR5*-Δ32 variant within the *CCR5* gene. When present the mutation confers resistance to HIV acquisition or slow progression in carriers (13,14). This finding has successfully been translated into virus entry inhibitor antiretrovirals (15,16). Other genes identified through candidate gene method include *HLA*-*A, HLA -B* and *HLA -C, CCR2, SDF1, IL10* (17,18), *CCL5* (RANTES), *KIR* genes, *TRIM5* and *APOBEC3G* (10–12).

The advent of genome-wide arrays uncovered several more loci associated with HIV-1 acquisition and progression. Genome-wide arrays did not require *a priori* knowledge of genomic region to be tested which rendered the method nearly unbiased (19). These methods once again bolstered the finding of *HLA-B* and *HLA-C* loci being the major determinants of HIV-1 control. Although the HLA region accounted for most of the variability in HIV-1 progression and control, GWAS of HIV-1 have uncovered new genes such as *ZNRD1* (20,21), *NOTCH4* (21,22), *C6orf48* (22,23) in European populations.

While the bulk of genome-wide association studies (GWAS) were carried out in European populations, few GWAS of African populations have been published to date (24–26). Of note, two novel variants within *HCG22* and *CCNG1* genes were found to be associated with progression and acquisition of HIV1 in an African (Botswana) population (26). We have previously discussed the results of GWAS of HIV-1 extensively here (27).

Genome-wide arrays revealed many variants of clinical significance to acquisition, control of HIV and progression (28). However, the method has limitations such as that 1) the tag-SNPs embedded in the arrays are not really causal, 2) the arrays harbour common variations so rarer variations can be missed and 3) due to higher diversity and lower linkage disequilibrium in African populations, the arrays are less robust in capturing variations in African populations (27–29).

Variants associated with HIV-1 in both candidate disease gene methods and GWAS accounted for just above 20% of the variability in the phenotypes of HIV (21,29). It has been proposed that this missing heritability of HIV-1 phenotypes may be hidden in rare variants among other factors (6,27). Whole genome (or exome) sequencing offers a more effective method to identify rare variants within African populations. We present here, whole genome sequencing (WGS) in a Southern African population of Botswana in efforts to pinpoint rare variants which may be of clinical significance to the acquisition of HIV-1.

## RESULTS

A total of 26,935 genes were used in the SKAT-O test, leading to a multiple test correction p-value cut-off of 1.86 × 10^−6^. Substructure among the HIV-1 positive/negative individuals was not observed, indicating similar genetic exposure among HIV-1 samples (30). Since no population has an entirely homogeneous genetic architecture, hidden relatedness and population structure have been accounted for in the current genetic association study.

### Aggregate rare-variant association of susceptibility to HIV-1 acquisition

No variant set reached statistical significance when controlling for possible confounding. With a SKAT univariate model including only HIV-1 status against the variant sets, we identified 194 variant sets with a p-value less than 0.01. The top effects included *Tet Methylcytosine Dioxygenase 1* (*TET1*) and 4 RNA genes which had not been reported previously (**Table 1**) in the GWAS of HIV-1.

**Table 1.**
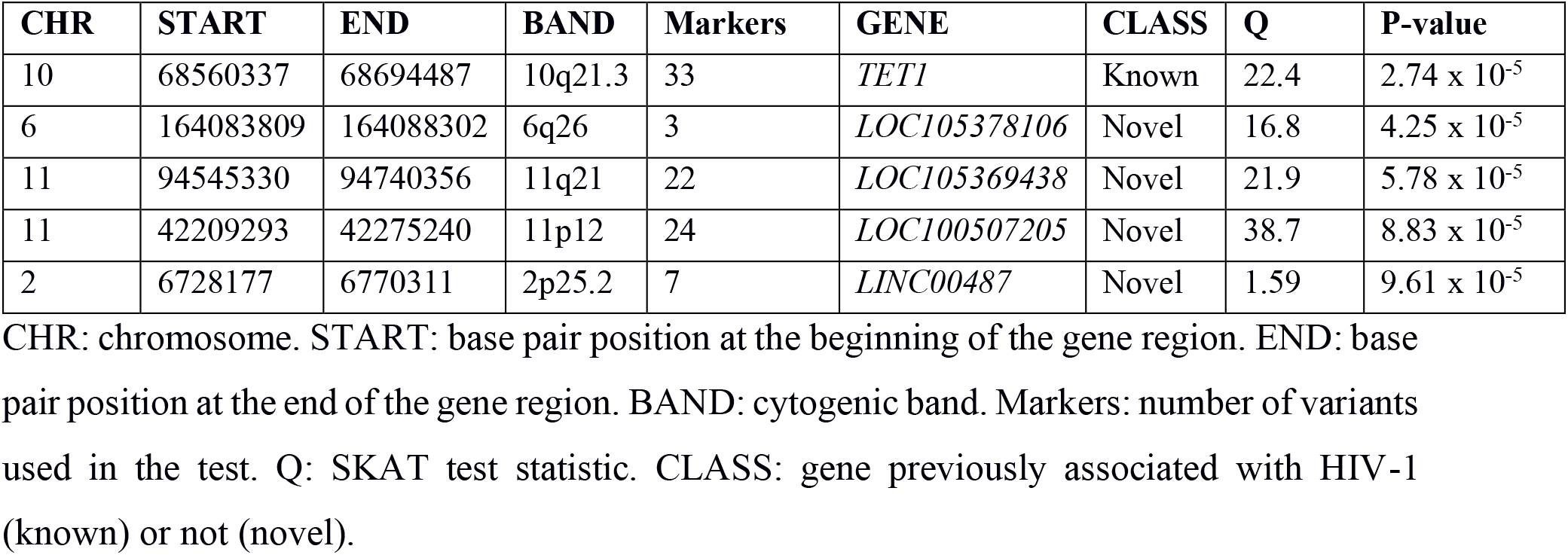
The strongest effects of the rare-variant association test of susceptibility to HIV-1 acquisition.

### 5.3.4.2 Aggregate rare-variant association of HIV-1 progression

Three sets of rare variants within the *Ankyrin Repeat Domain 39* (*ANKRD39*), *LOC105378523* and *General Transcription Factor IIIC Subunit 3* (*GTF3C3*) genes were statistically significant. Among the top 5 effects were the Metaxin (*MTX3*) and *Eukaryotic Translation Initiation Factor 3 subunit K* (*EIF3K*) genes, though not statistically significant. These genes have not been previously reported in the GWAS of HIV-1 (**Table 2, Table S1**). The λ_GC_ of the rare-variants was 0.99, which suggests that population structure and confounding was adequately controlled (**Figure 1**).

**Table 2.** The strongest effects of the rare-variant association of HIV-1 progression.

| CHR | START | END | BAND | Markers | GENE | CLASS | P-value |
| --- | --- | --- | --- | --- | --- | --- | --- |
| 2 | 96836611 | 96858016 | 2q11.2 | 2 | *ANKRD39* | Novel | $8.48 \times 10^{-8}$ |
| 10 | 121291608 | 121322945 | 10q26.12 | 3 | *LOC105378523* | Novel | $7.45 \times 10^{-7}$ |
| 2 | 196763035 | 196799725 | 2q33.1 | 7 | *GTF3C3* | Novel | $1.36 \times 10^{-6}$ |
| 5 | 79976716 | 79991262 | 5q14.1 | 1 | *MTX3* | Novel | $2.84 \times 10^{-6}$ |
| 19 | 38619082 | 38636955 | 19q13.2 | 1 | *EIF3K* | Novel | $2.92 \times 10^{-6}$ |

CHR: chromosome. START: base pair position at the beginning of the gene region. END: base pair position at the end of the gene region. BAND: cytogenic band. Markers: number of variants used in the test. Q: SKAT test statistic. CLASS: previously associated with HIV-1 (known) or not (novel) in a GWAS.

**Figure 1.**
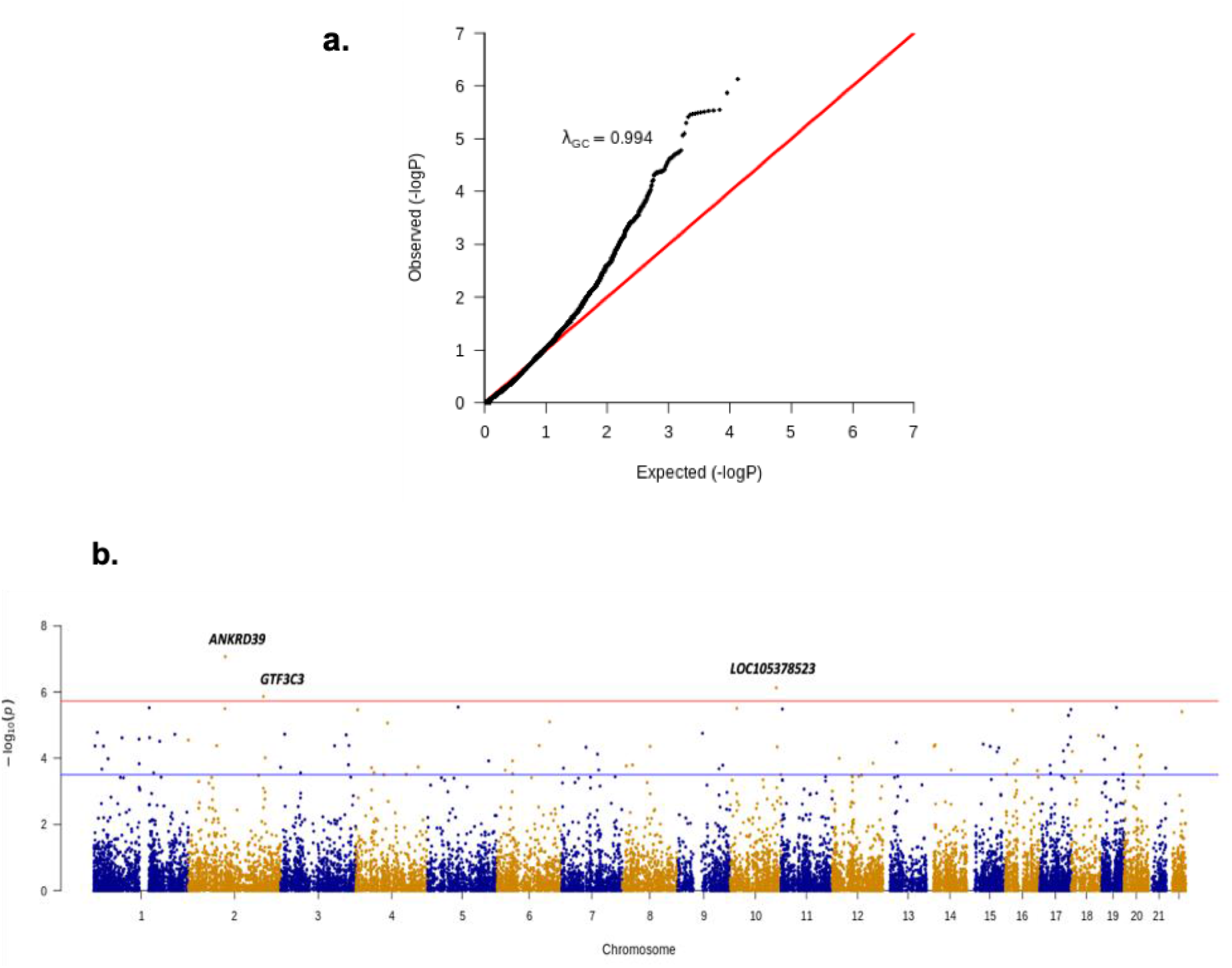
Quantile-quantile plot (with λ_GC_) and Manhattan plot of HIV-1 rare-variant association with HIV-1 progression. a. Quantile-quantile plot and λ_GC_ of HIV-1 rare-variant association with CD4+ T-cell counts showing -log_10_p-value of each variant is plotted against the expected null (the red line). b. Manhattan plot of HIV-1 rare-variant association with CD4+ T-cell counts showing -log_10_ p-value of each variant plotted against its genomic position. The red line is the -log_10_ p-value cut-off (1.86× 10^−6^).

### Gene-set enrichment of the strongest effects of genetic association of HIV-1 progression

In addition to the 5 candidate genes identified in common and rare-variant association of HIV-1 progression, 20 more related genes (grey circles without stripes) were retrieved through a gene-gene network (**Figure 2**). These genes were significantly (p-value < 0.05) enriched for the following biological processes: viral translation [(GO:0019081), p-value = 9.10 × 10^−16^], transcription from RNA polymerase III promoter [(GO:0006383), 9.46 × 10^−11^], and Cytoplasmic translational initiation [(GO:0002183), p-value = 2.51 × 10-6].

**Figure 2.**
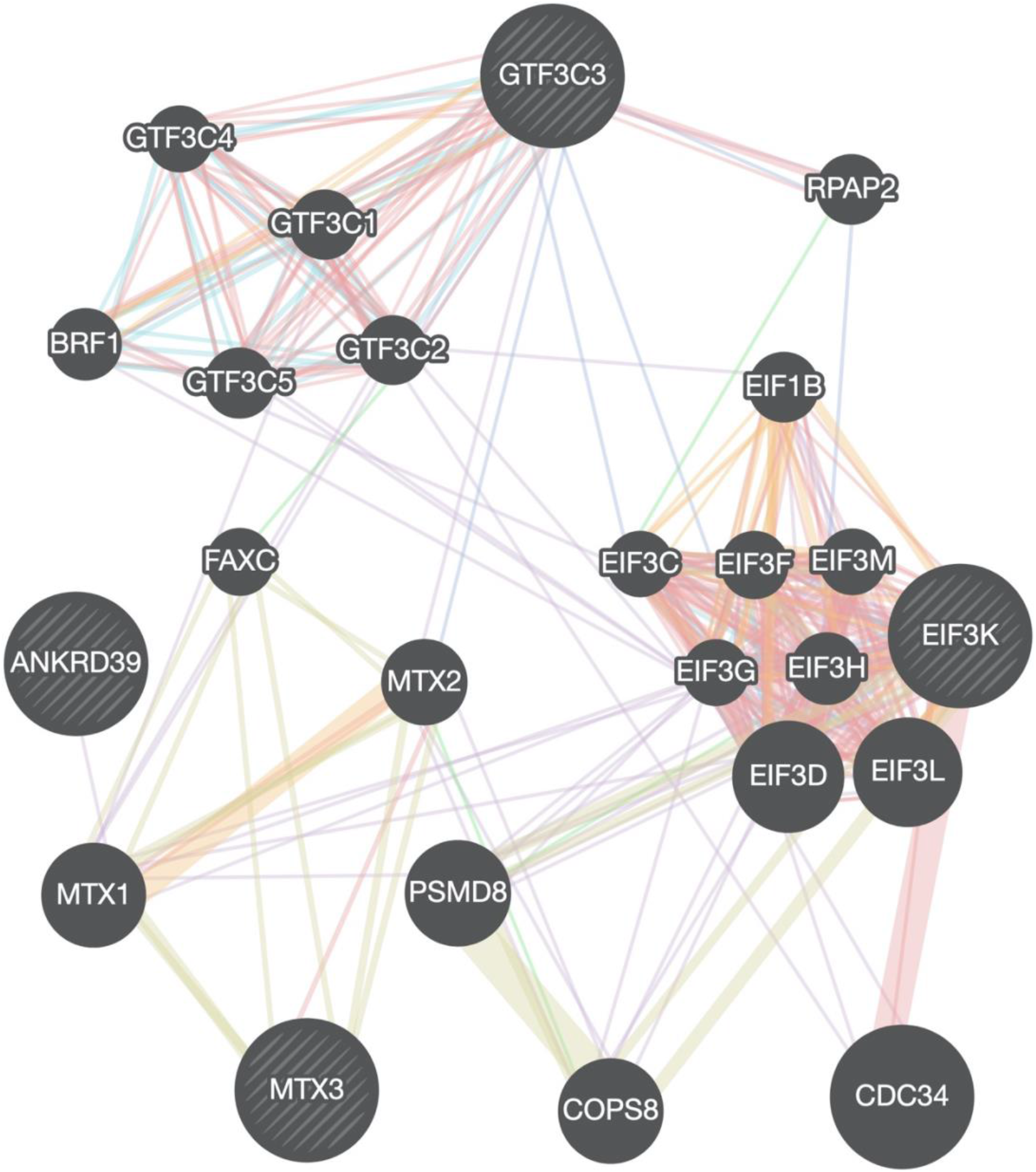
Gene interaction network of candidate genes identified from genetic association of HIV-1 progression. The different colours of branches represent how the genes are related; pink: physical interactions, purple: co-expression, orange: predicted, navy blue: co-localization, blue: Pathway, green: Genetic interactions, yellow: shared protein domains. Black and stripped nodes: genes provided as input into GeneMANIA (**Table 3**). Black only nodes: genes predicted by GeneMANIA to interact with the input list.

## DISCUSSION AND CONCLUSION

Our study is the first to evaluate the role of rare-variants in susceptibility to HIV-1 and progression to disease in Botswana using a larger sample size (n = 236), comparing to a previous study that had a cohort of 100 participants from Southern and Eastern African combined (at most 10 samples from Botswana) (33). In our study, the cumulative effects of three sets of novel rare variants within the *ANKRD39* (8.48 × 10^−8^), *LOC105378523* (7.45 × 10^−7^) and *GTF3C3* (1.36 × 10^−6^) genes were significantly associated with HIV-1 progression (**Table 2**).

**Table 3.** Enrichr gene-set enrichment of the candidate genes of HIV-1 progression.

|  | P-value | P-value<sub>adj</sub> | Database |
| --- | --- | --- | --- |
| **Gene Ontology** |  |  |  |
| Viral translation (GO:0019081) | 3.56 x 10<sup>-19</sup> | 9.10 x 10<sup>-16</sup> | Biological Process 2018 (31)<br>http://www.informatics.jax.org/ |
| Transcription from RNA polymerase III promoter (GO:0006383) | 1.30 x 10<sup>-13</sup> | 9.46 x 10<sup>-11</sup> |  |
| Cytoplasmic translational initiation (GO:0002183) | 4.43 x 10<sup>-9</sup> | 2.51 x 10<sup>-6</sup> |  |
| RNA polymerase III transcription factor complex (GO:0090576) | 1.21 x 10<sup>-18</sup> | 5.38 x 10<sup>-16</sup> | Cellular Component 2018 (31)<br>http://www.informatics.jax.org/ |
| Transcription factor TFIIC complex (GO:0000127) | 7.94 x 10<sup>-17</sup> | 1.77 x 10<sup>-14</sup> |  |
| Translation initiation factor activity (GO:0003743) | 1.57 x 10<sup>-37</sup> | 1.81 x 10<sup>-34</sup> | Molecular Function 2018 (31)<br>http://www.informatics.jax.org/ |
| RNA binding (GO:0003723) | 8.89 x 10<sup>-7</sup> | 3.41 x 10<sup>-4</sup> |  |
| **Pathways** |  |  |  |
| RNA polymerase III transcription initiation from type 2 promoter | 2.27 x 10<sup>-11</sup> | 4.29 x 10<sup>-9</sup> | BioPlanet 2019 (32) |
| Translation Factors | 2.84 x 10<sup>-22</sup> | 4.28 x 10<sup>-19</sup> | WikiPathways 2019 Human (Slenter et al, 2017) |
|  |  |  | BioCarta 2016<br>(http://www.biocarta.com) |

**P-value<sub>adj</sub>**: adjusted P-value.

The *ANKRD39* gene is not well characterized, however, it is a paralog of the *Cyclin Dependent Kinase Inhibitor 2D* (*CDKN2D*) that plays a role in regulation of cyclin and cell growth cycle (34). An artificial ankyrin repeat domain was found to negatively interfere with HIV-1 Gag assembly and budding (35). In addition, a common variant within a cyclin (*CCNG1*) gene was found to be associated with HIV-1 disease progression in a previous GWAS of Botswana (26). This supports the finding of the current study and possibly means that the rare variants within the *ANKRD39* gene might contribute to the control of DNA synthesis, cell division and consequently replication of HIV-1 in the host’s cells.

The *LOC105378523* gene is an uncharacterized lncRNA gene located within chromosome 10 (**Table 2**). The significant association of *LOC105378523* with CD4+ T-cell counts in the current study is expected as lncRNAs are known to be involved in regulation HIV replication, transcription and post-transcription, this makes them potential biomarkers for HIV-1 progression and targets for HIV treatment (36–39). Lastly, the *GTF3C3* gene (**Table 2**) encodes a transcription factor TFIIIC that is involved in transcription of transfer ribonucleic acid (tRNA) and directly binds to virus-associated RNA promoters (34). The transcription factor TFIIIC was observed to mediate HIV-1 transcription in HeLa and Jurkat T cells (40).

The functional analysis results accentuates that the candidate genes that potentially associate with HIV-1 progression affect RNA polymerase III transcription initiation, viral transcription and translation (**Table 3**). The RNA polymerase III transcription initiation pathway is represented by the *GTF3C* genes cluster (**Figure 2**). Upon integration into the human genome, HIV-1 RNA transcription is mediated by RNA polymerase II (41). Though the product of the *GTF3C3* gene has been linked to upregulation of HIV-1 RNA (40), in addition to mRNA synthesis, HIV-1 specific small nuclear RNAs can also be produced from RNA polymerase III promoters (42–44). HIV-1 hijacks the host translation machinery to facilitate its viral proteins. The human *EIF3* gene products (**Figure 2**) are some of the targets for optimal HIV-1 translation (45). These findings therefore suggest that the genes are involved in the regulation of HIV-1 transcription and translation which are markers of progression.

No variant set reached statistically significance in the rare-variant association test of susceptibility to HIV-1 acquisition. The strongest effects were within the *TET1* (p-value = 2.74 × 10^−5^), *LOC105378106* (p-value = 4.25 × 10^−5^), *LOC105369438* (p-value = 5.78 × 10^−5^), *LOC100507205* (p-value = 8.83 × 10^−5^) and *LINC00487* (p-value = 9.61 × 10^−5^) gene (**Table 1**). The product of the *TET1* gene is a demethylase that plays a role in DNA methylation and gene expression (34). TET (Ten-eleven translocation) family of proteins consists of three paralogs: TET1, TET2, and TET3 (46). It has been shown that the HIV-1 Vpr protein facilitates the degradation of TET-2 to promote HIV-1 replication (47,48). Furthermore, the HIV-1 Vpr protein also promotes HIV-1 Env processing and infectivity of macrophages which are known to be less susceptible to HIV-1 than CD4+ T-cells (48). This may imply that *TET1* gene plays a role in both HIV-1 progression and acquisition which supports the results of the current study that suggest that the *TET1* gene might be implicated in susceptibility to HIV-1 acquisition (**Table 1**). There has been a recent discovery of the association of a *CCR5* dependent lncRNA with susceptibility to HIV-1 (49), therefore, though the variant sets did not reach statistical significance, the identification of rare variant sets within lncRNAs in the current study (**Table 1**) cannot be entirely overlooked.

The cumulative effects of rare variants within the ANKRD39, LOC105378523 and GTF3C3 were found to be significantly associated with HIV-1 progression. To the best of our knowledge, none of these genes have been previously associated with HIV-1 through GWAS. Identifying these candidate disease genes warrants their further investigation as a potential target in HIV-1 pathogenesis in African populations. Although additional data was not available at the time of analysis, in future studies increasing the sample-size could improve the results of susceptibility to HIV-1 acquisition. There is also need for a development of statistical test rare-variant association that is designed to handle longitudinal phenotypic data as the current optimized rare-variant association tests are limited in this regard.

## MATERIALS AND METHODS

### Ethical approval

This study is part of a bigger protocol titled “Host Genetics of HIV-1 Subtype C Infection, Progression and Treatment in Africa/GWAS on determinants of HIV-1 Subtype C Infection” conducted by Botswana Harvard AIDS Institute Partnership. Ethics approval was obtained according to The Declaration of Helsinki. All participants consented to participate in the study. Institutional Review Board (IRB) approval was obtained for these samples from Botswana Ministry of Health and Wellness - Health Research Development Committee (HRDC) & Harvard School of Public Health IRB (reference number: HPDME 13/18/1) and the University of Cape Town - Human Research Ethics Committee (HREC reference number: 316/2019).

### Selection of study participants and sample preparation

The study participants and sample preparation have been described previous in our previous work (30). Additionally, the HIV-1 negative participants enrolled in the current study were those that self-reported that they were highly exposed to HIV-1 and required HIV-1 testing in the Tshedimoso study. The Tshedimoso study sought to identify acute and recent HIV-1C infection in Botswana. These participants remained negative for the duration of the study (50).

### Whole genome sequence Data Description

Quality assessment was performed on paired-end WGS (minimum of 30X depth) in FASTQ format (51) using FastQC (52). Low-quality sequence bases and adapters were trimmed using Trimmomatic with default parameters (53). The sequencing reads were aligned to the GRCh38 human reference genome using Burrows-Wheeler Aligner (BWA-MEM) (54,55) and post-alignment quality control including adding of read groups, marking duplicates, fix mating and recalibration of base quality scores was performed using Picard tools, SAMtools (56) and Genome Analysis Toolkit (57).

We performed population joint calling (58,59) using two different methods to leverage the quality and accuracy of our results: GATK HaplotypeCaller (57,60) and BCFtools (56). The variant call format (VCF) dataset was filtered using VCFTOOLS (61), GATK Variant Quality Score Recalibration and BCFtools. The specific filtering parameters employed for both call-sets have been detailed (30). Downstream analyses were performed with GATK call-set and BCFtools call-set used as a validation set. Further quality control as required prior genetic association tests was performed using PLINK (62). Variants that had genotype missingness and MAF of 5% or more, and those that deviated from Hardy-Weinberg Equilibrium (HWE p > 1.0 × 10^−5^) were filtered out.

### Rare-variant association tests

To account for sample size and rare variants, (that standard GWAS could have missed or not reached the genome-wide significant level) we leverage possible small effects by aggregating SNPs effects at gene level with an optimal unified sequence kernel association test (SKAT-O) (63,64). This test combined burden and variance-component analyses using the SKAT package (65) in order to appropriately discriminate 265 HIV-1 positive and 125 HIV-1 negative individuals. SKAT(-O) has been optimized to control for type I error and increase power for small sample sizes. Moreover, the SKAT_NULL_emmaX() module was used with kinship relatedness matrix to adjust for genetic relatedness and population structure within the data. A p-value of 0.05/genes was considered significant; where genes is the number of genes tested in the model.

For HIV-1 progression, we regressed CD4+ T-cell counts over several covariates (**Table S2**) using lmer function in the R lme4 package (66). We then used the regression coefficients (slopes) of each individual to infer CD4+ T-cell changes over a period of at least 18 months (236 young women). A number of models were tested to select confounding variables and interacting factors (Table S1). As expected, CD4+ T-cell counts were significantly correlating with the presence of highly active antiretroviral therapy (HAART), this justifying the inclusion of HAART as a covariate to model CD4+ T-cell changes over time. The CD4+ T-cell slopes were used as a quantitative phenotype for genetic subsequent association tests.

### Pathways enrichment analysis and gene-gene interactions

Functional analysis was performed to elucidate variant effect mechanisms by identifying putatively affected biological processes and pathways. The genes identified in the rare-variant association of HIV-1 progression were subjected to gene-set enrichment analysis using GeneMANIA (67) and Enrichr (68) bioinformatics tools.

## Data Availability

Requests to access the sequence data should be addressed to

## ACKNOWLEDGEMENTS

This work was supported through the sub-Saharan African Network for TB/HIV Research Excellence (SANTHE), a DELTAS Africa Initiative [grant # DEL-15-006]. The DELTAS Africa Initiative is an independent funding scheme of the African Academy of Sciences (AAS)’s Alliance for Accelerating Excellence in Science in Africa (AESA) and supported by the New Partnership for Africa’s Development Planning and Coordinating Agency (NEPAD Agency) with funding from the Wellcome Trust [grant # 107752/Z/15/Z] and the UK government. The views expressed in this publication are those of the authors and not necessarily those of AAS, NEPAD Agency, Wellcome Trust, or the UK government. The authors would also like to thank the National Research Foundation of South Africa for funding (NRF) [grant # RA171111285157/119056].

## AUTHORS CONTRIBUTIONS

PKT and ERC conceived and structured the content of the manuscript. PKT conducted data analysis and result interpretation. PKT drafted the manuscript. PKT, ERC, DDM, WTC, CD, MML, VN, SJO, ME and SG edited the manuscript.

